# Usage and awareness of antiviral medications for COVID-19

**DOI:** 10.1101/2022.04.21.22274155

**Authors:** N Kojima, JD Klausner

## Abstract

We surveyed people that recently tested positive for SARS-CoV-2 to assess the frequency and correlates of early treatment seeking behavior. Among high risk respondents, 66.0% were aware of treatment for COVID-19 and 36.3% had sought treatment, however only 1.7% reported use of an antiviral for SARS-CoV-2 infection. More public outreach is needed to raise awareness of the benefits of treatment for COVID-19.

## Main Text

Coronavirus disease 2019 (COVID-19), caused by severe acute respiratory syndrome coronavirus 2 (SARS-CoV-2), continues to cause severe disease, hospitalization, and death. There are effective means to treat SARS-CoV-2 infection [1-4], however is it not clear how often those who would benefit most use monoclonal antibodies, molnupiavir, and nirmatrelvir. We surveyed users of a national SARS-CoV-2 testing company to assess treatment awareness, treatment seeking behavior, and use of antiviral medications for those with a positive test result.

We recruited adults (18 years or older) who had tested positive for SARS-CoV-2 by a reverse transcriptase polymerase chain reaction (RT-PCR) assay at a large clinical laboratory (Curative, San Dimas, CA). To be eligible, individuals had to have a positive RT-PCR test result within 7 days of enrollment. All surveys were anonymous and voluntary. We collected data on age, assess treatment awareness, treatment seeking behavior, and use of antiviral medications for COVID-19. Descriptive statistics were calculated on StataSE (StataCorp, College Station, TX). Advarra deemed the study IRB exempt (Pro00059961).

There were 1,159 respondents during March 2022. Among individuals aged 65 years and older, 66.0% were aware of treatment for COVID-19 and 36.3% had sought treatment, however only 1.7% reported use of an antiviral for SARS-CoV-2 infection (Figure). Rates were similar among younger individuals.

**Figure.**
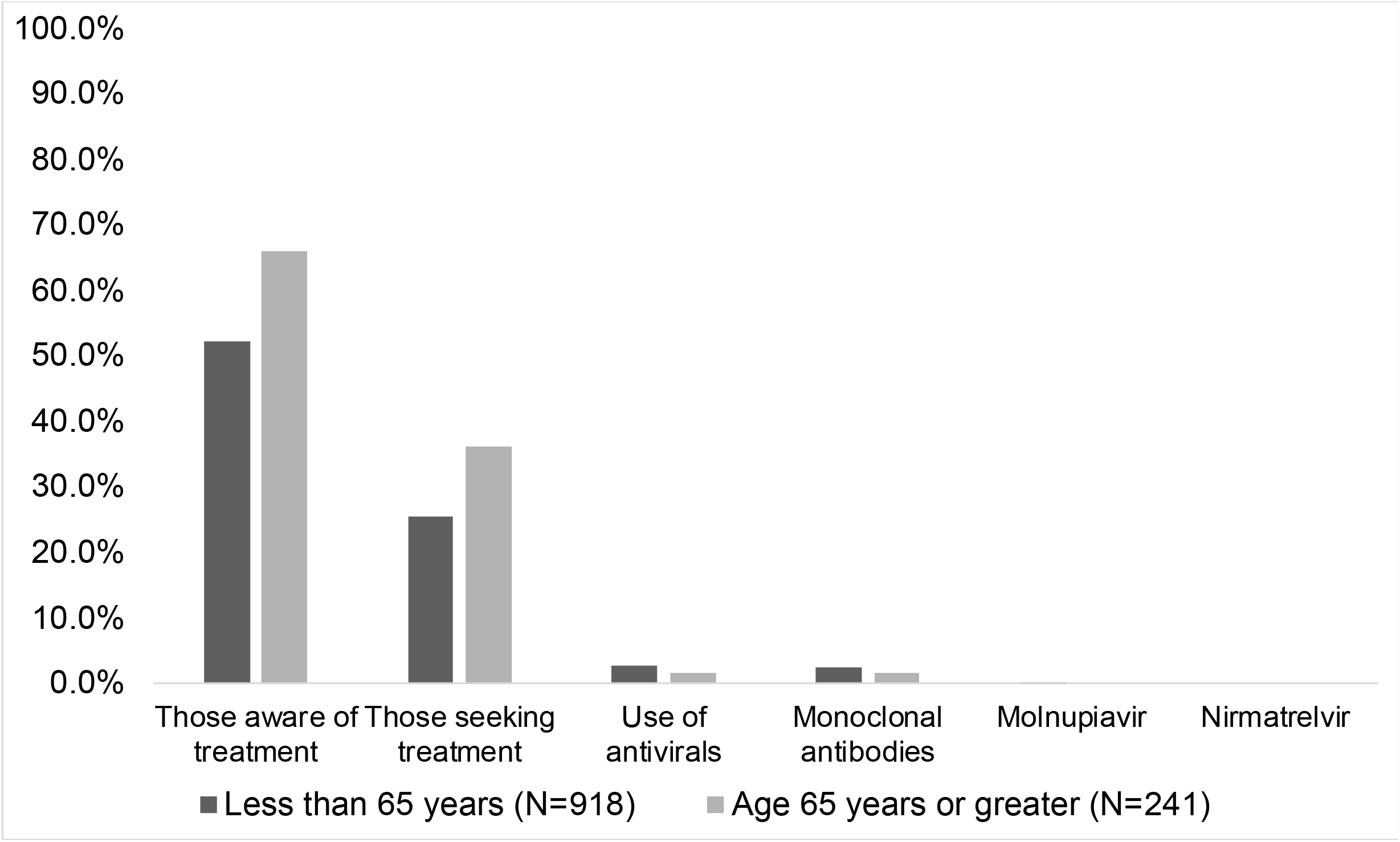
Treatment awareness, treatment seeking behavior, and use of antiviral medications among individuals 65 years and older versus individuals younger than 65 years that tested positive for SARS-CoV-2 infection in 37 States in the United States from 1 March 2022 to 30 March 2022

Due to the timing of the survey, infections were most likely caused by either the Omicron variant of SARS-CoV-2 or a sublineage of the Omicron variant, which might be less severe in those with prior immunity [5]. Our findings demonstrate that increased awareness and use of antivirals among those at risk for severe disease due to COVID-19 are urgently needed among individuals that test positive for SARS-CoV-2.

## Data Availability

All data produced in the present study are available upon reasonable request to the authors.

## Declarations

### Declaration of competing interests

NK is a consultant for Curative. JDK is an independent consultant and serves as the Medical Director of Curative.

### Funding

Curative Inc. and by a gift to the Keck School of Medicine of the University of Southern California by the W.M. Keck Foundation.

## Acknowledgements

To the participants that donated their time.

## Notes

### Author Declarations

Advarra deemed the study IRB exempt (Pro00059961).

### Summary of Updates

Corrected incorrect dates in figure title

